# Acute Interleukin 17A and Cortisol Responses to the Maastricht Acute Stress Test in Healthy Adults

**DOI:** 10.64898/2026.07.23.26358762

**Authors:** Benedict Herhaus, Lara Jürgens, Markus Moehler, Rupert Conrad, Katja Petrowski

## Abstract

Interleukin-17A (IL-17A) has been implicated in stress- and pain-related inflammation, yet evidence for acute IL-17A stress response in humans remains scarce. This study examined whether the Maastricht Acute Stress Test (MAST) induces changes in circulating IL-17A and how IL-17A dynamics relate to cortisol and subjective stress/pain. Forty-six healthy adults (mean age: 30.50 ± 12.86 years; 54% female) completed the standardized MAST. Serum IL- 17A and cortisol were assessed repeatedly across baseline and recovery. Stress- and pain-related ratings (VAS) and trait questionnaires (e.g., chronic stress, psychological distress, resilience) were completed. There was a significant increase over time in IL-17A and cortisol following the MAST, with an 36 % increase of IL-17A level measured at 105 min post-stressor and cortisol peaking at 10 min post-stressor. IL-17A indices (baseline and AUCi) showed no meaningful associations with cortisol, subjective stress/pain, or psychological traits, whereas cortisol response (AUCi) correlated positively with perceived acute stress and pain. Overall, the MAST produced clear IL-17A and cortisol responses that appeared dissociable in healthy individuals. Future studies should include broader inflammatory panels and clinical or high-stress samples to clarify conditions under which IL-17A covaries with HPA-axis activity and subjective experience.

## 1. Introduction

Interleukin-17 (IL-17) is a pro-inflammatory cytokine family that plays a central role in host defence and immune-mediated tissue inflammation (Pappu et al., 2011). While the IL-17 family comprises six different cytokines, IL-17A is the prototypical one, discovered more than two decades ago with broaden impact on immune processes (McGeachy et al., 2019). Predominantly produced by T helper 17 (Th17) cells, but also by innate lymphoid cells, γδ T cells, and neutrophils, IL-17A acts at the interface of innate and adaptive immunity (Chung et al., 2021). It promotes the recruitment and activation of neutrophils and induces the expression of cytokines, chemokines, and antimicrobial peptides (AMPs) in epithelial and stromal cells (Pappu et al., 2011). Therefore, IL-17A signalling is essential for protection against extracellular pathogens (Qian et al., 2010). While tightly regulated, its dysregulation contributes to chronic inflammation and has been implicated in the pathogenesis of several autoimmune and inflammatory diseases (Zhu and Qian, 2012). IL-17A signalling is closely intertwined with hypothalamic–pituitary–adrenal (HPA) axis activity. Glucocorticoids such as cortisol exert potent immunoregulatory effects by repressing pro-inflammatory transcriptional programmes and can thereby constrain cytokine networks that support Th17/IL-17 responses (De Bosscher et al., 2003). Conversely, IL-17A–associated inflammation has been linked to reduced glucocorticoid responsiveness (e.g., via impaired anti-inflammatory signalling), suggesting bidirectional crosstalk between cortisol output and IL-17A pathways (Zijlstra et al., 2012).

Recently, IL-17A has gained increasing attention as a central immune mediator linking psychological stress to neuroinflammatory processes and pain sensitization (Jiang et al., 2022; Mamun-or-Rashid et al., 2024). Preclinical evidence suggests that stress can modulate IL-17A expression in the brain, supporting a stress–inflammation link (Dragasevic et al., 2025; Liu et al., 2012). In the cumulative mild stress (CPMS) mouse model, Kim et al. (2021) reported that CPMS leads to long-lasting anxiety- and depression-like behaviors, accompanied by increased IL-17A and microglial activation in the hippocampus, amygdala, and prefrontal cortex. In humans, chronic psychosocial stress and mental disorders have been associated with elevated IL-17A activity (Mamun-or-Rashid et al., 2024; Moore et al., 2019). For example, Mamun-or-Rashid et al. (2024) reported increased IL-17A levels in patients with generalized anxiety disorder and positive associations with symptom severity, while Moore et al. (2019) found IL-17A to correlate positively with multiple stress measures in pregnant women. Notably, evidence for acute IL-17A stress response in humans is limited. In a laboratory study using the PASAT — often considered a comparatively mild, primarily cognitive stressor relative to more robust paradigms such as the TSST or MAST — Maydych et al. (2018) found no significant change in salivary IL-17A following stress exposure.

Beyond its role in stress-related neuroinflammation, IL-17A is also increasingly implicated in pain processing, as it can promote peripheral and central sensitization by amplifying pro-inflammatory signaling and glial activation (Jiang et al., 2022). IL-17A activity has been reported to be elevated across several chronic pain conditions (e.g., neuropathic, inflammatory, and cancer pain), and pain itself may act as an additional stressor (Jiang et al., 2022). In rheumatoid arthritis, serum IL-17A was significantly higher in individuals with anxiety, and IL-17A levels correlated with anxiety severity even after adjusting for pain and disease activity (Liu et al., 2012). Experimental work using an intermittent cold stress (ICS) model in mice further supports a mechanistic link between stress, IL-17A elevation, and pain: ICS increased IL-17A in plasma and pain-relevant brain regions and was accompanied by fibromyalgia-like mechanical and thermal hypersensitivity (Yeh et al., 2024). Notably, electroacupuncture reduced IL-17A-related inflammatory signaling and attenuated stress-induced pain behaviors, supporting IL-17A as a mediator in stress-driven pain amplification (Yeh et al., 2024). In humans, repeated cold pressor test (CPT) exposure likewise elicited salivary immune responses, although salivary IL-17A was largely below the assay’s detection limit, limiting conclusions about IL-17A pain response in this paradigm (Larra et al., 2023). Taken together, preclinical and clinical findings suggest that IL-17A may be involved in stress-related neuroimmune alterations and pain sensitization. However, human evidence on acute IL-17A response remains scarce and methodologically limited, as existing studies have largely relied on salivary measurements and comparatively mild stress or pain protocols, with inconsistent detectability. To address this gap, the present study examines whether a robust standardized acute stressor—the Maastricht Acute Stress Test (MAST)—elicits measurable changes in circulating IL-17A. Therefore, the current aim was to study the IL-17A stress response under the standardized stress protocol of the Maastricht Acute Stress Test (MAST), which combines psychosocial stress with cold pressor pain (Smeets et al., 2012). Given evidence that IL-17A is linked to stress-related psychopathology and is elevated across chronic pain conditions (Jiang et al., 2022; Liu et al., 2012; Mamun-or-Rashid et al., 2024; Moore et al., 2019), we hypothesized that higher chronic stress and chronic pain perceptions would be associated with higher baseline (resting) IL-17A levels (Hypothesis 1). Because acute laboratory stress reliably increases several circulating inflammatory cytokines in humans (e.g., IL-6, IL-1β, TNF-α; (Marsland et al., 2017)), we hypothesized that IL-17A may show an acute increase in blood following the MAST (Hypothesis 2). A negative cortisol–IL-17A association was expected, as cortisol has predominantly anti-inflammatory effects via glucocorticoid receptor–mediated repression of pro-inflammatory signaling (De Bosscher et al., 2003). Consequently, we hypothesized that healthy individuals showing stronger cortisol stress responses may display a more constrained IL-17A response (Hypothesis 3).

## 2. Materials and Methods

### 2.1 Study participants

Forty-six participants (men and women) were recruited through electronic tendering (e- tendering) and via notice boards at Johannes Gutenberg University Mainz. Eligibility was assessed using a web-based self-report screening questionnaire in which inclusion and exclusion criteria were reviewed. Individuals were eligible if they were aged 18–65 years. Participants were excluded in case of acute or chronic medical illness, endocrinological disorders, obesity (BMI ≥ 30 kg/m²), psychiatric or other mental disorders, medication or substance use, current intake of psychotropic medication or corticosteroid preparations, major stressful life events within the past six months, or smoking more than ten cigarettes per day. The participants’ mean age was 30.50 ± 12.86 years, and their mean body mass index (BMI) was 23.40 ± 2.92 kg/m². Table 1 provides a comprehensive overview of participant characteristics, including demographic information and psychological status. The study protocol received approval from the local Ethics Committee of the Landesärztekammer Rheinland-Pfalz, Germany (No. 2023-16934).

**Table 1.** Characteristics of the participants.

|  |  |
| --- | --- |
| Age (years), M (SD) | 30.50 (12.86) |
| Females, n (%) | 25 (54) |
| BMI (kg/m<sup>2</sup>), M (SD) | 23.40 (2.92) |
| Systolic blood pressure, mmHg, M (SD) | 126 (16) |
| Diastolic blood pressure, mmHg, M (SD) | 80 (12) |
| RS-13, M (SD) | 74.78 (9.65) |
| Pain Resilience scale, M (SD) | 46.36 (6.67) |
| PHQ stress scale, M (SD) | 3.85 (3.30) |
| BSI-18, M (SD) | 5.11 (6.60) |
| TICS-9, M (SD) | 9.84 (6.25) |

Abbreviations: BSI, Brief Symptom Inventory; M, mean; PHQ, Patient Health Questionnaire; RS, Resilience scale; SD, standard deviation; SSS-8, Somatic Symptom Scale; TICS, Trier Inventar for Chronic Stress.

**Table 2.** Fixed effects from the mixed model examining the interaction between IL-17A as well cortisol and Time-Points during the Maastricht Acute Stress Test.

| IL-17A as dependent variable |  |  |  |  |  |
| --- | --- | --- | --- | --- | --- |
| Effect | Estimate | SE | df | t | p |
| (Intercept) | 1.160 | .14 | 45 | 8.16 | <□.001 |
| +60 - -1 | .143 | .05 | 135 | 2.76 | .007 |
| +75 - -1 | .252 | .05 | 135 | 4.85 | < .001 |
| +105 - -1 | .364 | .05 | 135 | 7.01 | < .001 |
| Cortisol as dependent variable |  |  |  |  |  |
| Effect | Estimate | SE | df | t | p |
| (Intercept) | 71.321 | 3.78 | 44 | 18.86 | <□.001 |
| +1 - -1 | 17.932 | 3.10 | 350 | 5.78 | <□.001 |
| +10 - -1 | 25.660 | 3.10 | 350 | 8.28 | <□.001 |
| +20 - -1 | 16.488 | 3.10 | 350 | 5.32 | <□.001 |
| +30 - -1 | 8.276 | 3.10 | 350 | 2.67 | 0.008 |
| +45 - -1 | -0.656 | 3.10 | 350 | -0.21 | 0.833 |
| +60 - -1 | -6.841 | 3.10 | 350 | -2.21 | 0.028 |
| +75 - -1 | -13.008 | 3.12 | 350 | -4.19 | <□.001 |
| +105 - -1 | -19.075 | 3.12 | 350 | -6.11 | <□.001 |

### 2.2 Procedures

The laboratory visit took place between 2:00 p.m. and 5:00 p.m. Participants underwent the Maastricht Acute Stress Test (MAST) according to Smeets et al. (2012) and were instructed to refrain from eating, drinking, and smoking prior to and throughout the approximately two-hour visit. To minimize procedure-related effects on endocrine and immune outcomes (e.g., pain-related increases in cortisol and IL-17A), an intravenous cannula was inserted 45 min before the first blood draw. The protocol started with a 15-min baseline period during the first blood sample (-1 min) was collected. Participants then completed the 15-min MAST. The initial 5 min consisted of standardized instructions and task familiarization. During the subsequent 10-min stress induction, the MAST combined a physical and a psychosocial/cognitive stressor: participants repeatedly immersed their hand in cold water (cold pressor component) while simultaneously performing a mentally demanding arithmetic task under time pressure and social-evaluative monitoring. Thus, both the cold-water immersion and the arithmetic task were administered for a total of 10 min. After each cold-water immersion, they rated pain on a visual analogue scale VAS, and the mean of all pain ratings was calculated. Immediately after the stress condition, participants rated subjective stress on a VAS. Following completion of the MAST, participants remained in a supine position for 105 min. During this recovery period, blood samples were collected at +1, +10, +20, +30, +45, +60, +75, and +105 min.

### 2.3 Psychological and Clinical Measures

The PHQ Stress Scale (PHQ-Stress; (Löwe et al., 2004)) was administered to assess psychosocial stressors and the degree to which participants felt burdened by them over the past four weeks. The scale comprises 10 items rated on a 3-point scale (0–2), with higher sum scores indicating greater psychosocial stress. General psychological resilience was measured with the Resilience Scale–13 (RS-13; (Leppert et al., 2008)), a 13-item self-report instrument rated on a 7-point Likert scale (1–7). Item scores are summed, with higher total scores reflecting higher resilience. Pain-specific resilience was assessed using the Pain Resilience Scale (PRS; (Slepian et al., 2016)), a 14-item questionnaire capturing two dimensions— cognitive/affective positivity and behavioral perseverance—in the context of intense or prolonged pain. Items are typically rated on a 5-point scale (0–4), with higher scores indicating greater pain resilience. Chronic stress was assessed with the short version of the Trier Inventory for Chronic Stress (TICS) described by Petrowski et al. (2019). This short form contains 9 items representing the nine theoretical stress domains of the TICS and refers to experiences during the previous three months, rated on a 5-point frequency scale (0–4); higher scores indicate higher chronic stress. Current psychological distress was measured with the Brief Symptom Inventory–18 (BSI-18; (Franke, 2000)), which assesses symptoms across somatization, depression, and anxiety (6 items each) over the past seven days on a 5- point scale (0–4).

### 2.4 Blood analytics

After intervention, blood samples were collected in serum gel monovettes (S-Monovette® 9 ml Z, Sarstedt, Nümbrecht, Germany) to measure serum IL-17A levels. The collected monovettes were left at room temperature for 30 min to facilitate blood coagulation. Following coagulation, the serum monovettes were centrifuged for 10 min at 2500 g and 20 °C. Serum IL-17A levels were assessed using a Human High Sensitivity Interleukin 17A (HS IL-17A) ELISA (Invitrogen, Thermo Fisher Scientific, USA). Serum cortisol concentrations were quantified using a commercially available enzyme-linked immunosorbent assay (ELISA) kit (IBL International GmbH, Germany).

### 2.5 Statistical Analysis

The effect of the MAST on IL-17 was analyzed using a mixed model, with interaction of the measurement points (-1, +60, +75, +105) as a fixed effect. In view of a statistically meaningful sample size, the optimum statistical sample size was calculated with the G*power program (version: 3.1.9.2.) (Faul et al., 2007). Drawing on the meta-analysis by Marsland et al. (2017), which examined inflammatory cytokine response to acute laboratory stressors, cytokine stress response in healthy adults was found to be small-to-moderate in magnitude. A power analysis showed that for a small-to-moderate effect size (Coheńs f =.175), four measurement points, significance level of p =.05 and power of 80% (1-ß =.80), a total sample size of n = 46 participants for ANOVA-repeated measures (between factors) is needed. We tested 49 participants, but three participants were excluded from analyses regarding IL-17 concentration, since their measured values were more than three standard deviations from the mean in all measurement points of a condition (Schlotz, 2013). The data were analyzed according to the normality of distributions and were, in case of not normally distributed data, subjected to logarithm naturalis transformations. In addition, we calculated the area under the response curve with respect to increase (AUCi) for IL-17 and cortisol to index stress-induced changes over time (Pruessner et al., 2003). These AUCi indices were then used in Pearson correlation analyses to examine whether endocrine and inflammatory response (AUCi) and baseline concentrations were associated with MAST pain and stress ratings as well as questionnaire-based trait measures (pain resilience, PHQ stress, BSI-18, and TICS-9). All statistical analyses were conducted using RStudio (Version 2025.05.1) and R (Version 4.5.1) using the packages lme4 (Version 1.1-34), lmerTest (Version 3.1-3), emmeans (Version 1.11.2), car (Version 3.1.3), dplyr (1.1.3), and effectsize (Version 1.0.1). Visualization of the data was performed with Prism v.8.3.0 (GraphPad Software).

## 3. Results

### IL-17A response to acute stress

Figure 1 (A, B) presents the IL-17A concentration over four measurement points during the MAST. The highest increase of IL-17A concentrations (5.x pg/ml ± SD) was measured 105 min post-stressor compared to baseline (4.x pg/ml ± SD). In total, the 46 healthy participants showed a 36% increase relative to baseline in the IL-17A concentration. In the fixed-effects omnibus test, time-points had a significant effect on IL-17A, *F*(3, 135) = 17.90, *p* < .001 (Satterthwaite degrees of freedom). The estimated IL-17A level at the reference time point (−1; intercept) was *b* = 1.160 (*SE* = 0.142, *df* = 45), *t* = 8.16, *p* < .001. Relative to −1, IL-17A was significantly higher at +60 (*b* = 0.143, *SE* = 0.052, *df* = 135), *t* = 2.76, *p* = .007, at +75 (*b* = 0.252, *SE* = 0.052, *df* = 135), *t* = 4.85, *p* < .001, and at +105 (*b* = 0.364, *SE* = 0.052, *df* = 135), *t* = 7.01, *p* < .001.

**Figure 1.**
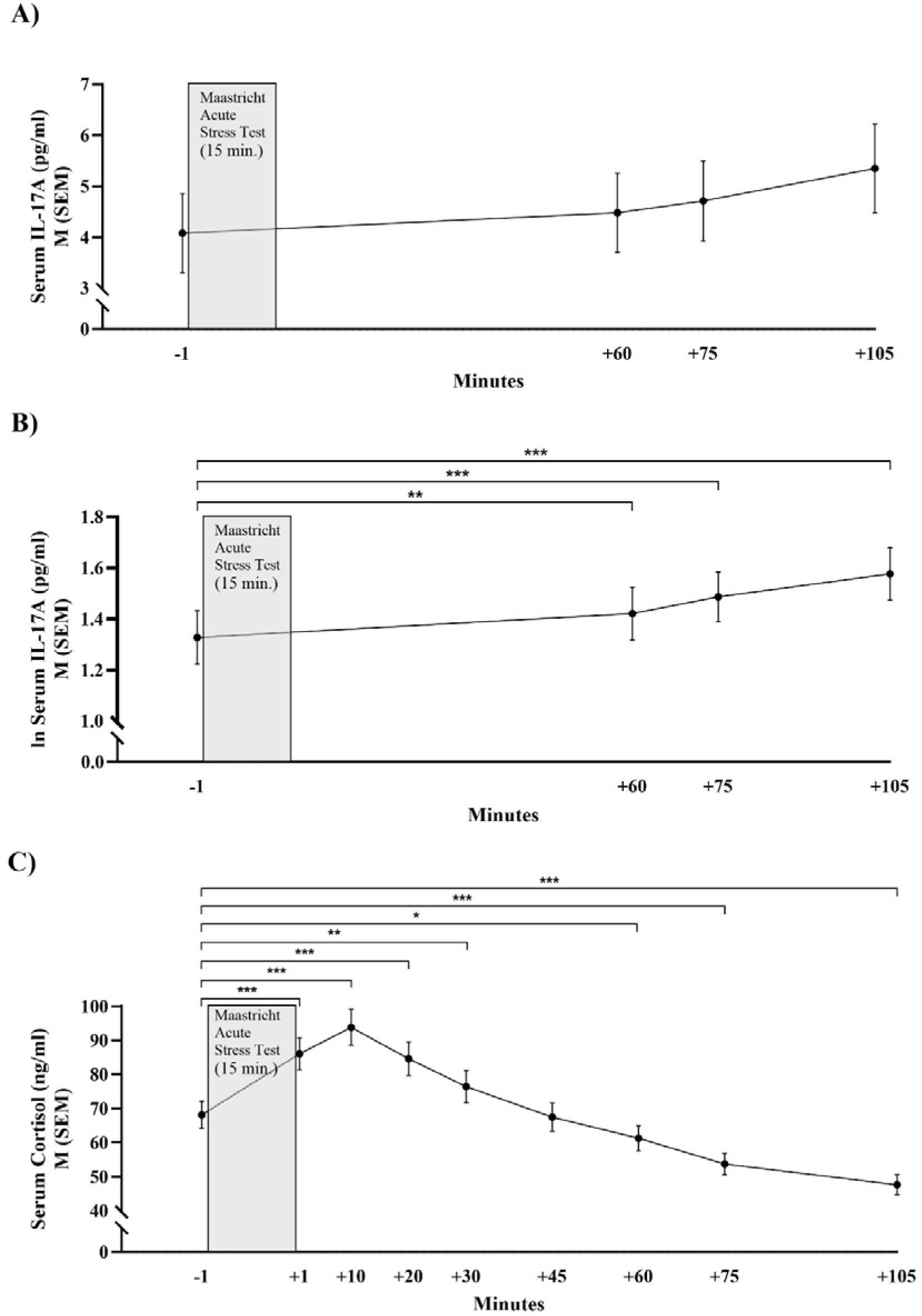
Serum IL-17A concentration (A: raw data, B:logarithm naturalis transformed data) and Cortisol (C) in healthy men and women during the Maastricht Acute Stress Test. * < 0.05, ** < 0.01, *** < 0.001

### Cortisol response to acute stress

Figure 1 (C) presents the cortisol concentration over nine measurement points during the MAST. The 46 healthy participants showed a 43% increase 10 min post-stressor relative to baseline in the cortisol concentration. In the fixed-effects omnibus test, time-points exerted a significant effect on cortisol, *F*(8, 350) = 46.50, *p* < .001 (Satterthwaite degrees of freedom). The estimated cortisol level at the reference time point (−1; intercept) was *b* = 71.321 (*SE* = 3.78, *df* = 44), *t* = 18.86, *p* < .001. Relative to −1, cortisol was significantly higher at +1 (*b* = 17.932, *SE* = 3.10, *df* = 350), *t* = 5.78, *p* < .001, +10 (*b* = 25.660, *SE* = 3.10, *df* = 350), *t* = 8.28, *p* < .001, +20 (*b* = 16.488, *SE* = 3.10, *df* = 350), *t* = 5.32, *p* < .001, and +30 (*b* = 8.276, *SE* = 3.10, *df* = 350), *t* = 2.67, *p* = .008. Cortisol did not differ from −1 at +45 (*b* = −0.656, *SE* = 3.10, *df* = 350), *t* = −0.21, *p* = .833. Thereafter, cortisol was significantly lower than −1 at +60 (*b* = −6.841, *SE* = 3.10, *df* = 350), *t* = −2.21, *p* = .028, +75 (*b* = −13.008, *SE* = 3.12, *df* = 350), *t* = −4.19, *p* < .001, and +105 (*b* = −19.075, *SE* = 3.12, *df* = 350), *t* = −6.11, *p* < .001.

### Associations between psychological and IL-17A and cortisol measures

Across the primary biomarkers and MAST outcomes, IL-17A showed no meaningful associations (see Table 3). Neither baseline lnIL-17A nor IL-17A AUC_I_ correlated significantly with baseline cortisol, cortisol AUCI, pain/stress VAS, or psychological trait measures (all *ps* ≥ .256). In contrast, cortisol indices were related to MAST responses. Cortisol AUC_I_ was positively associated with both pain VAS (*r* = .35, *p* = .020) and stress VAS (*r* = .49, *p* < .001).

**Table 3.** Pearson’s correlations (r) between psychological and IL-17A and cortisol measures.

|  | Baseline<br>IL-17 | AUC<sub>I</sub><br>IL-17 | Baseline<br>Cortisol | AUC<sub>I</sub><br>Cortisol | Pain VAS<br>after<br>MAST | Stress<br>VAS after<br>MAST | Pain<br>Resilience<br>scale | PHQ<br>stress<br>scale | BSI-18 | TICS-9 |
| --- | --- | --- | --- | --- | --- | --- | --- | --- | --- | --- |
| Baseline<br>IL-17A<sup>†</sup> | 1.00 |  |  |  |  |  |  |  |  |  |
| AUC<sub>I</sub><br>IL-17A | .09<br>*p* = .573 | 1.00 |  |  |  |  |  |  |  |  |
| Baseline<br>Cortisol | .07<br>*p* = .646 | .02<br>*p* = .913 | 1.00 |  |  |  |  |  |  |  |
| AUC<sub>I</sub><br>Cortisol | .00<br>*p* = .991 | .07<br>*p* = .644 | -.41<br>*p* = .005 | 1.00 |  |  |  |  |  |  |
| Pain VAS<br>after<br>MAST | .12<br>*p* = .432 | -.14<br>*p* = .358 | -.32<br>*p* = .035 | .35<br>*p* = .020 | 1.00 |  |  |  |  |  |
| Stress VAS<br>after<br>MAST | -.03<br>*p* = .865 | .01<br>*p* = .963 | -.12<br>*p* = .453 | .49<br>*p* < .001 | .44<br>*p* = .002 | 1.00 |  |  |  |  |
| Pain<br>Resilience<br>scale | .07<br>*p* = .654 | -.04<br>*p* = .804 | .01<br>*p* = .974 | -.14<br>*p* = .366 | .12<br>*p* = .436 | -.09<br>*p* = .546 | 1.00 |  |  |  |
| PHQ stress<br>scale | .06<br>*p* = .697 | .07<br>*p* = .632 | .26<br>*p* = .087 | .28<br>*p* = .066 | .08<br>*p* = .616 | .11<br>*p* = .451 | -.47<br>*p* = .001 | 1.00 |  |  |
| BSI-18 | -.04<br>*p* = .787 | .12<br>*p* = .421 | .34<br>*p* = .025 | .18<br>*p* = .250 | .07<br>*p* = .639 | .12<br>*p* = .424 | -.41<br>*p* = .004 | .76<br>*p* < .001 | 1.00 |  |
| TICS-9 | .17<br>*p* = .256 | .04<br>*p* = .783 | .08<br>*p* = .596 | .14<br>*p* = .355 | .04<br>*p* = .781 | .13<br>*p* = .408 | -.50<br>*p* < .001 | .66<br>*p* < .001 | .70<br>*p* < .001 | 1.00 |

Data are presented as coefficient, *p* valuesAUC<sub>I</sub>, area under the curve with respect to increase; BSI, Brief Symptom Inventory; PHQ, Patient Health Questionnaire; TICS Trier Inventory of Chronic Stress; VAS, Visual Analog Scale. †, logarithm naturalis transformed data

## 4. Discussion

This is the first study to investigate IL-17A response to the standardized acute stressor MAST and its temporal dynamics in relation to cortisol. As expected, the MAST elicited a robust increase in IL-17A (∼36% above baseline). Cortisol also rose markedly (∼43%), showing a significant increase during the immediate post-stressor phase, a return to baseline thereafter, and a subsequent decline below baseline consistent with pronounced recovery/overshoot. Whereas IL-17A indices (baseline and AUC_I_) showed no meaningful associations with cortisol, cortisol response (AUC_I_) was positively related to perceived stress and pain during the acute stressor.

The stress-induced increase in IL-17A observed following the MAST contrasts with prior null findings following comparatively mild, primarily cognitive stressors such as the PASAT (Maydych et al., 2018), suggesting that IL-17A may be stress-responsive when the challenge is sufficiently intense. This interpretation is supported by the robust cortisol rise, indicating effective HPA-axis activation, and aligns with meta-analytic evidence that acute laboratory stress reliably elevates circulating inflammatory cytokines (e.g., IL-6, IL-1β, TNF-α; (Marsland et al., 2017)). One proposed mechanism is stress-hormone–driven immune cell mobilization and trafficking (catecholamines and glucocorticoids), which can transiently increase circulating inflammatory signals (Dhabhar et al., 2012). Notably, the temporal dynamics of IL-17A are likely to differ from those of cortisol, as glucocorticoid responses reflect rapid ACTH-driven adrenal steroidogenesis, whereas IL-17A depends on immune-cell activation and transcriptional regulation of cytokine expression, particularly in Th17 cells (Capone and Volpe, 2020; Turcu and Auchus, 2015).

Converging evidence from animal models points to a functional link between adrenal stress hormones and IL-17A activity: in inflammatory mouse paradigms, endogenous glucocorticoids suppress IL-17A expression/release, whereas experimental removal of adrenal hormones is associated with markedly elevated IL-17A (Bosmann et al., 2013). Complementary human-focused work on stress–immune interactions further suggests that while cortisol typically constrains pro-inflammatory signaling, chronic stress and glucocorticoid resistance may weaken this inhibitory control, allowing IL-17A–related activity to remain elevated (Nunez et al., 2025). In contrast to this proposed neuroendocrine– immune crosstalk, no meaningful associations were observed between IL-17A indices and cortisol measures.

Several factors may explain the absence of detectable IL-17A–cortisol associations in this healthy sample. First, cytokine–HPA coupling is often modest and highly time-dependent; meta-analytic work highlights substantial heterogeneity in stress-induced inflammatory responses as a function of baseline inflammatory tone, sampling schedules (e.g., latency to peak cytokine response), assay sensitivity, and participant characteristics (Marsland et al., 2017; Steptoe et al., 2007). Second, many immunoregulatory effects of glucocorticoids are mediated via glucocorticoid receptor–dependent genomic mechanisms (i.e., changes in transcription and de novo protein synthesis) that typically unfold over hours rather than minutes, which may weaken straightforward, linear associations with short-term fluctuations in circulating IL-17A in low-inflammatory healthy samples (Ayroldi et al., 2012; Kadmiel and Cidlowski, 2013). In line with this, our analysis was limited to a 105-min post-stress window and captured only the initial increase in IL-17A, while potential peak levels and subsequent declines may have occurred outside this observation period. Third, IL-17A regulation appears strongly context-dependent: experimental inflammatory models show suppression of IL-17 family expression by adrenal hormones, but these effects are most evident under pronounced immune activation and may not translate directly to acute psychosocial stress in healthy humans (Bosmann et al., 2013).

Despite prior evidence linking IL-17A to chronic psychosocial stress, anxiety-related symptom burden, and chronic pain states (Jiang et al., 2022; Mamun-or-Rashid et al., 2024; Moore et al., 2019), our findings showed no meaningful associations between IL-17A indices (baseline or AUC_I_) and psychological trait measures, subjective stress/pain ratings. One possible explanation may be the use of a healthy, non-clinical sample with comparatively low levels and restricted variance in chronic stress and pain, limiting the detectability of IL-17A correlates. Thus, IL-17A may reflect longer-term inflammatory dysregulation rather than subtle differences in stress perception among otherwise healthy individuals. In contrast, the HPA-axis marker showed clear links to subjective experience: cortisol output (AUCi) correlated positively with acute perceived stress and pain, suggesting that individual differences in perceived pain intensity and stressfulness of the task were closely mirrored in cortisol response. This pattern is consistent with evidence that stressors high in uncontrollability and social-evaluative threat elicit stronger cortisol responses, and that correspondence between perceived stress and cortisol is most apparent under sufficiently robust laboratory stress conditions (Campbell and Ehlert, 2012; Dickerson and Kemeny, 2004; Gaab et al., 2005).

A major strength of this study is the use of the standardized, well-validated acute stress test MAST and the frequent sampling of serum IL-17A and cortisol to accurately measure stress-induced dynamics, including peaks and recovery processes in a sample with a well-balanced distribution of sex and age. However, several limitations should be noted. The exclusive focus on healthy, highly screened participants likely restricted variance in stress/pain burden and basal inflammation, which can attenuate biomarker–psychological associations (Lakes, 2013). Moreover, the sample size was powered for within-person cytokine response based on meta-analytic estimates, whereas biomarker–trait correlations were secondary and may have been underpowered for small effects (Marsland et al., 2017). Finally, inflammatory profiling was limited to IL-17A; adding broader cytokine panels (e.g., IL-6, IL-1β, TNF-α, CRP) and complementary Th17/IL-17A pathway markers (e.g., IL-17A/IL-23–related gene expression or immune cell phenotyping) would help delineate mechanisms of stress-induced immune activation (Kaymaz and Beikler, 2019; Marsland et al., 2017).

In conclusion, the MAST elicited robust cortisol and IL-17A responses, but only cortisol response showed meaningful links to subjective stress and pain. These findings suggest that acute psychosocial stress reliably engages HPA-axis activity, whereas IL-17 changes may be less tightly coupled to psychological stress experience in healthy individuals. Future studies should incorporate broader inflammatory panels, clinical or high-stress samples, and extended post-stressor follow-up sampling to clarify when IL-17A covaries with HPA-axis activity and subjective experience and to capture the full temporal dynamics of acute stress– induced IL-17A responses, including potential peak levels and subsequent declines.

## Data Availability

All data produced in the present study are available upon reasonable request to the authors

## Acknowledgements

We thank the medical PhD students for assisting with data collection.

## Conflict of interests

The authors have no conflict of interest to disclose.

## Source of Funding

This study was funded by the European Union project ‘RELEVIUM’ (Project-number: 101057821).

## Declaration of Generative AI and AI-assisted technologies in the writing process

During the preparation of this work the author(s) used ChatGPT in order to improve readability and language. After using this tool/service, the author(s) reviewed and edited the content as needed and take(s) full responsibility for the content of the publication.

## Notes

### Competing Interest Statement

The authors have declared no competing interest.

### Author Declarations

The Ethics Committee of the State Medical Association of Rhineland-Palatinate (Ethik-Kommission bei der Landesaerztekammer Rheinland-Pfalz) gave ethical approval for this work (approval no. 2023-16934). All participants provided written informed consent before participation.

